# Assessment of Bias in Estimates of Sexual Network Degree using Prospective Cohort Data

**DOI:** 10.1101/19003830

**Authors:** Stephen Uong, Eli S. Rosenberg, Steven M. Goodreau, Nicole Luisi, Patrick Sullivan, Samuel M. Jenness

## Abstract

**Background:** Sexual network degree, a count of ongoing partnerships, plays a critical role in the transmission dynamics of human immunodeficiency virus (HIV) and other sexually transmitted infections (STI). Researchers often quantify degree using self-reported cross-sectional data on the day of survey, which may result in bias because of uncertainty about future sexual activity.

**Methods:** We evaluated the bias of a cross-sectional degree measure with a prospective cohort study of men who have sex with men (MSM). At baseline, we asked men about whether recent sexual partnerships were ongoing. We confirmed the true, ongoing status of those partnerships at baseline at follow-up. With logistic regression, we estimated the partnership-level predictors of baseline measure accuracy. With Poisson regression, we estimated the longitudinally confirmed degree as a function of baseline predicted degree.

**Results:** Across partnership types, the baseline ongoing status measure was 70% accurate, with higher negative predictive value (91%) than positive predictive value (39%). Partnership exclusivity and racial pairing were associated with higher accuracy. Baseline degree generally overestimated confirmed degree. Bias, or number of ongoing partners different than predicted at baseline, was -0.28 overall, ranging from -1.91 to -0.41 for MSM with any ongoing partnerships at baseline. Comparing MSM of the same baseline degree, the level of bias was stronger for black compared to white MSM, and for younger compared to older MSM.

**Conclusions:** Research studies may overestimate degree when it is quantified cross-sectionally. Adjustment and structured sensitivity analyses may account for bias in studies of HIV or STI prevention interventions.

## INTRODUCTION

An estimated 1.1 million persons in the United States were living with human immunodeficiency virus (HIV), with 40,000 new diagnoses occurring in 2017.^1^ Despite substantial progress in the development of new tools for HIV prevention, including preexposure prophylaxis (PrEP), incidence has stabilized in the past half-decade.^2^ Men who have sex with men (MSM) accounted for 66% of new HIV diagnoses but less than 5% of the population in 2017,^3^ and new cases have increased among several marginalized MSM subgroups such as young black and Hispanic MSM.^2^ Meeting HIV prevention targets, such as the newly established *Ending the HIV Epidemic Plan* calling for a 90% reduction in HIV incidence by 2030,^4^ will require sustained efforts to understand the drivers of HIV infection among MSM, the causes of disparities, and optimal methods for targeting prevention tools.

Networks of sexual partnerships have long been a focus of HIV research.^5^ A network framework addresses the ongoing challenge that individual-level behavior and biology, by themselves, do not sufficiently explain the size of the HIV epidemic.^6^ Sexual partner concurrency, defined by having two or more ongoing partnerships, has been identified as a central explanatory cause for the shape of the HIV epidemic in Sub-Saharan Africa and the large sex differentials in HIV prevalence there.^7,8^ Concurrency amplifies the speed of HIV transmission across the population, compared to serial monogamy with the same number of cumulative partners.^9^ Concurrency is a binary categorization of momentary network degree (hereafter, degree), which is the number of ongoing partners at any point in time. Sexual networks may be characterized by a range of features, but degree, assortative mixing, and partnership duration are three important measures for HIV transmission.^10^ Recent network studies among MSM have characterized the interacting effects of these three features on the trajectory of HIV incidence.^11,12^

Network degree and duration are typically measured through cross-sectional study designs given the difficulty in longitudinal assessment. Although the time period for quantifying degree in these designs has been debated,^13^ one preferred approach is a measure on the day of study. Use of day-of-study degree is preferred because it allows for joint estimation of degree and duration following common statistical assumptions.^14^ Duration may be based on partnership start and end dates with the latter censored for ongoing partnerships. A challenge with the day-of-study degree measure, however, is that it requires study participants to predict whether partnerships will continue. Because of uncertainty in that prediction, measured degree may be a biased estimator of true degree. If the ongoing status of partnerships is systematically overpredicted, estimates for network degree and partnership duration would be biased upward.

Assessment of bias in estimates of network degree is uncommon because of the validation data needed. Linked partnership studies have evaluated agreement in degree within sexual dyads, and retrospective studies have evaluated temporal changes in degree over time.^15,16^ But no studies have assessed the accuracy of a day-of-study degree measure. Such bias assessment would require prospective data in which persons would be asked at follow-up to confirm whether partnerships reported as ongoing at baseline were truly ongoing, and whether those reported as not ongoing were truly not. The bias in the cross-sectional only degree measurement would likely have important heterogeneity by demographic characteristics. If black MSM misreport degree more than white MSM, for example, the explanatory power of degree for research on racial disparities is weakened.^17^ Bias also likely varies by partnership characteristics, such as partnership type; ongoing status is less often known in casual partnerships.^18^ However, the epidemiologic impact of this bias for casual partnerships may be less than for main partnerships because the former are shorter in duration.

Validation of degree measures has important implications for both mathematical modeling research and applied prevention activities. Biases in these network parameters could result in incorrect projections for mathematical models that simulate disease transmission dynamics or intervention effects.^10^ Network features are often used, implicitly or explicitly, in many HIV/STI prevention activities such as partner notification to prevent within-partnership reinfection,^20^ and in novel HIV/STI prevention interventions that target broader features of the sexual network structure.^21^

In this study, we use prospective cohort data on reported partnerships to quantify how accurately participants predicted the continuing status of their partnerships, as well as the total number of ongoing partnerships (degree). To maximally inform HIV/STI prevention and mathematical modeling activities, we evaluated how accuracy, as well as negative and positive predictive values, varied across definitions of baseline degree, and by key factors. Our broader goal was to estimate the level of potential bias in degree to generate bias adjustment factors for future HIV/STI prevention research and interventions.

## METHODS

### Study Design

This analysis used data from Involvement, a prospective cohort study in 2010–2014, to investigate multilevel factors for HIV risk among black and white MSM in the Atlanta metropolitan area.^22^ Study procedures included a standardized survey measuring behavioral, biologic, and sexual network attributes hypothesized to influence HIV risk. We recruited study participants through structured time–location sampling of sites where MSM congregated in Atlanta, supplemented with web-based recruitment. We purposefully selected locations and time periods to increase enrollment of black MSM to ensure a balanced cohort.

Enrollment eligibility criteria were male sex, age between 18 and 39, non-Hispanic black or white race, residence in the Atlanta Metropolitan Statistical Area, at least one male sex partner within the past three months, and not being in a mutually monogamous (either the participant or their partners were not exclusive) relationship. Overall, 560 of the 803 participants screened as HIV-negative at baseline and enrolled into the cohort for follow-up. At each follow-up, participants received HIV and bacterial STI screening and completed additional behavioral surveys. Previous reports have described the sampling, recruitment, and enrollment protocols in further detail.^22,23^ The Emory University Institutional Review Board approved this study.

### Measures

Our analyses included measures at the baseline and Month 6 (M6) follow-up visits. At baseline, participants reported on up to their five most recent partners over the prior 6 months. Specific question wording for relevant measures from these two surveys are included in eFigure 1. Questions included whether participants considered these partnerships as ongoing (those in which participant expected to have sexual contact again). Participants could report yes, no, or don’t know to these questions. At the M6 visit, we asked participants again about the same partners they reported on at baseline, including whether any sexual activity occurred after the baseline visit. The ongoing status at baseline of those partnerships reported could be confirmed with this M6 data in that way. The surveys included additional questions on the duration and frequency of sexual acts within the partnership, reported on previously,^23^ but those measures were not needed in this analysis as we considered partnerships only by their ongoing status (i.e., partnerships were not stratified in this analysis based on the subsequent duration or number of acts after baseline).

**Figure.**
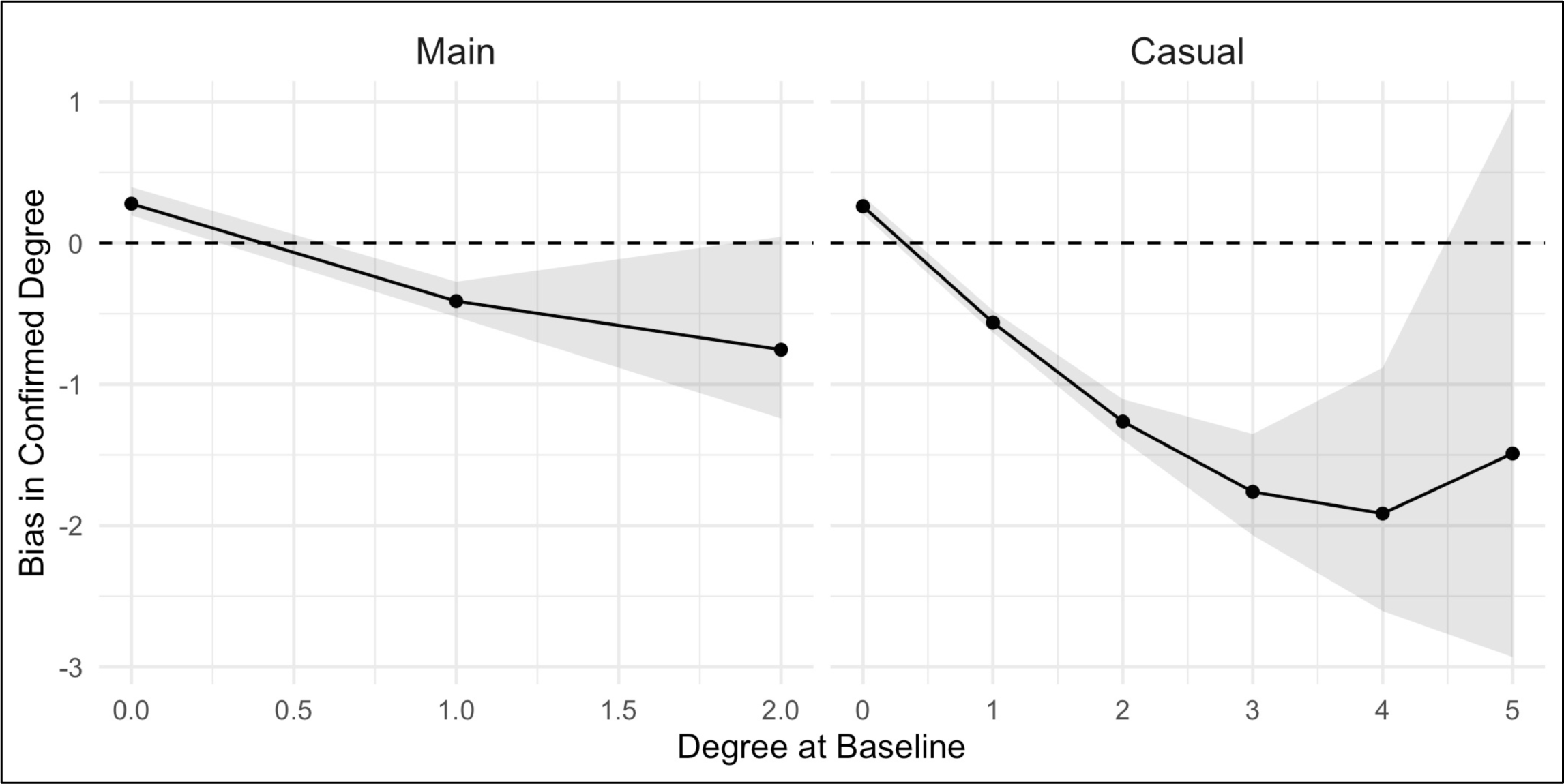
Difference between confirmed and baseline degree as a function of baseline degree, stratified by partnership type. Dashed horizontal line at 0 displays value where there is no difference between baseline and confirmed degree. Predictions below the dashed line indicate that baseline degree overestimated confirmed degree. Lines/dots display the point estimates and grey polygons display the 95% confidence intervals around the estimates.

Degree refers to the sum of all ongoing partnerships at baseline. *Baseline Degree* is the number of ongoing partnerships reported on baseline data only, and *Confirmed Degree* is the baseline degree confirmed with M6 data. Confirmed degree would be lower than baseline degree if partnerships categorized as ongoing at baseline were not truly ongoing upon reevaluation at M6.

Confirmed degree would be higher than baseline degree if partnerships categorized as not ongoing at baseline were truly ongoing upon reevaluation at M6. We define the bias in baseline degree as the difference between confirmed degree and baseline degree.

For this analysis, we excluded partnerships with women, those missing ongoing partnership status at baseline or month 6 follow-up, and partners with an unknown confirmed degree measure. We evaluated several individual predictors for degree agreement and bias, including age, race, and number of male sex partners. Partner-level covariates for predictions of accuracy for specific relationships included partnership type, frequency of sexual contact, race combination, age homophily, perceived concordant HIV status, and agreement about having outside sexual partnerships. Partnerships were categorized as main (primary with repeated sexual contacts), casual (non-primary but repeated sexual contacts), and one-time as of baseline. Participants could project that partnerships that were one-time as of baseline could continue in the same way as partnerships with multiple acts (casual or main partnerships). Participants could misclassify different relationships as ongoing or not, while still correctly estimating their overall momentary degree, if the numbers misclassified in each direction were equal.

### Statistical Analyses

To evaluate how well baseline ongoing status was predicted by confirmed M6 data, we calculated positive predictive value (proportion of partnerships predicted at baseline as ongoing that were confirmed ongoing), negative predictive value (proportion of partnerships predicted at baseline as not ongoing that were confirmed not ongoing), and accuracy (proportion of partnerships for which the baseline and M6 ongoing status values matched). We defined unknown baseline status as either not ongoing (unknown=no) or missing (unknown=missing; observations were dropped) to evaluate sensitivity to missing data assumptions.

We then used partnership-level logistic regression models (analytic unit: partnerships) to estimate predictors of accuracy of baseline ongoing status within partnerships. Partnership-level predictors of interest were race and age homophily, frequency of sex, perceived partner HIV status, and agreement about outside sexual partnerships. Sandwich variance estimators for robust standard errors were used to calculate 95% confidence intervals to account for clustering of partnerships within participants. Finally, we estimated the association between baseline and confirmed degree with individual-level Poisson regression models (analytic unit: participants) for main and casual partnership types. Baseline degree was the primary predictor, confirmed degree was the outcome, and race and age were included as hypothesized confounders. These models were used to estimate the direction and magnitude of the degree bias. All analyses were conducted using R 3.5.3.

## RESULTS

Compared to all MSM enrolled in the Involvement cohort, the analytic sample (N = 469) were of a similar race and age composition (Table 1). With respect to partnership characteristics, partnerships in the analytic sample (N = 1397) were more likely to be main and casual than one-time partnerships compared to those reported by the full cohort (N = 1758). Most partnerships (94%) were within the same race and nearly 40% were perceived to be HIV discordant (meaning the reported partner was HIV-infected, given that the cohort enrolled only HIV-uninfected MSM at baseline) or unknown (meaning the partner’s HIV status was unknown).

**Table 1.** Individual-Level and Partnership-Level Characteristics of Enrolled Cohort and Analytic Sample in a Study of Black and White Men Who Have Sex with Men, Atlanta, 2010–2014

| Characteristic | Enrolled Cohort |  | Analytic Sample |  |
| --- | --- | --- | --- | --- |
| *Individual* | *N = 560* |  | *N = 469* |  |
| **Age (n, %)** |  |  |  |  |
| 18–19 | 40 | 7.1 | 32 | 6.8 |
| 20–24 | 189 | 33.8 | 150 | 32.0 |
| 25–29 | 165 | 29.4 | 140 | 29.9 |
| 30–39 | 166 | 29.6 | 147 | 31.3 |
| **Race (n, %)** |  |  |  |  |
| Black | 257 | 45.9 | 205 | 43.7 |
| White | 303 | 54.1 | 264 | 56.3 |
| **Sexual Partners in Past 6 Months (mean, standard deviation, median)** |  |  |  |  |
| Total Partners | 5.7, 8.3, 5 |  | 5.8, 8.9, 5 |  |
| Condomless Partners | 1.9, 3.4, 1 |  | 1.8, 3.5, 1 |  |
| *Partnership* | *N = 1758* |  | *N = 1397* |  |
| **Partnership Type (n, %)** |  |  |  |  |
| Main | 267 | 15.3 | 253 | 18.1 |
| Casual | 533 | 30.5 | 494 | 35.4 |
| One-Time | 945 | 54.2 | 650 | 46.5 |
| **Racial Composition (n, %)** |  |  |  |  |
| Black–Black | 557 | 39.6 | 449 | 39.0 |
| Black–White | 95 | 6.8 | 73 | 6.4 |
| White–White | 754 | 53.6 | 628 | 54.6 |
| **Age Homophily (mean, standard deviation)** |  |  |  |  |
| Absolute Difference in Years | 8.1, 1.9 |  | 8.1, 1.7 |  |
| **Perceived Concordant HIV Status (n, %)** |  |  |  |  |
| Concordant | 965 | 57.9 | 814 | 60.6 |
| Discordant/Unknown | 702 | 42.1 | 530 | 39.4 |
| **Coital Frequency in Past 6 Months (mean, standard deviation, median)** |  |  |  |  |
| Total Frequency | 12.6, 18.4, 4 |  | 12.7, 18.5, 4 |  |
| **Partnership Agreement about Outside Sex (n, %)** |  |  |  |  |
| No Agreement | 565 | 71.6 | 527 | 71.6 |
| No Outside Sex | 112 | 14.2 | 103 | 14.0 |
| Outside Sex with Conditions | 43 | 5.45 | 40 | 5.4 |
| Outside Sex without Conditions | 69 | 8.8 | 66 | 9.0 |

The analytic sample includes only men with same-sex partnerships, no missingness in report of baseline and 6-month degree, and no unknown responses for 6-month degree.

Table 2 shows the accuracy of the baseline reported ongoing status, as confirmed with the M6 data as the gold standard. The sum of partnerships with baseline and confirmed ongoing statuses of each partnership equals the baseline and confirmed degree for each participant, respectively; marginal degree distributions are reported in eTable 1. Overall, there was largely consistency between the two measures (accuracy > 50%), with variation by partnership type and classification of unknown status partnerships. Limited to known baseline ongoing status (yes/no responses only, where baseline unknown responses were set to missing), accuracy was 63% overall, highest in main partnerships (69%), and lowest in casual partnerships (58%). When classifying unknown status partnerships as not ongoing, accuracy overall improved to 70% and increased to 72% in one-time partnerships. The row percentages may be interpreted as positive and negative predictive values when unknown ongoing partnerships were classified as missing. These show that the accuracy is driven by the negative predictive values (86%–92%), which was much higher than positive predictive values (19%–58%). The negative predictive values are reduced when classifying unknown ongoing partnerships as not ongoing (the positive predictive values would not change with this different classification of these partnerships).

**Table 2.** Accuracy of Baseline Reported Ongoing Status as Confirmed at 6-Month Follow-Up in a Study of Black and White Men Who Have Sex with Men, Atlanta, 2010–2014.

| Partnership Type | Baseline Ongoing | Confirmed Ongoing |  | Baseline Accuracy |  | Negative Predictive Value |
| --- | --- | --- | --- | --- | --- | --- |
|  |  | Yes | No | Unknown = Missing | Unknown = No | Unknown = No |
| Main (N = 253) | Yes | 73 (58%) | 54 (43%) | 69% | 67% | 77% |
|  | No | 11 (14%) | 69 (86%) |  |  |  |
|  | Unknown | 18 (39%) | 28 (61%) |  |  |  |
| Casual (N = 494) | Yes | 92 (42%) | 125 (58%) | 58% | 67% | 86% |
|  | No | 9 (9%) | 94 (91%) |  |  |  |
|  | Unknown | 29 (17%) | 145 (83%) |  |  |  |
| One-Time (N = 650) | Yes | 29 (19%) | 127 (81%) | 64% | 72% | 89% |
|  | No | 21 (8%) | 239 (92%) |  |  |  |
|  | Unknown | 32 (14%) | 202 (86%) |  |  |  |
| Overall (N = 1397) | Yes | 194 (39%) | 306 (61%) | 63% | 70% | 87% |
|  | No | 41 (9%) | 402 (91%) |  |  |  |
|  | Unknown | 79 (17%) | 375 (83%) |  |  |  |

Table 3 presents the results of logistic regression models for partnership-level predictors of accuracy of the baseline ongoing status. The categorization of accuracy was the same as in Table 2. Accuracy of baseline degree measures was higher in partnerships with smaller age gaps, white– white racial composition, main partnership type (compared to casual partnership type), and agreement about partnership exclusivity. When unknown status partnerships were coded as not ongoing, each 5-year increase in absolute age gap was associated with 9% lower odds of accuracy. That association was strengthened (OR = 0.87 versus 0.91) when treating unknown partnerships as missing. White–white partnerships across types were 1.4 times more accurate compared to black– black partnerships, and the effect was larger in main white–white partnerships compared to casual white–white partnerships. Partnership agreements about exclusivity with respect to outside partners were associated with accuracy, but this was driven by main partnerships. Perceived HIV status and coital frequency had minimal and inconsistent relationships to accuracy.

**Table 3.** Partner-Level Logistic Regression of Accuracy of Baseline Reported Ongoing Status as Confirmed at 6-Month Follow-Up, in a Study of Black and White Men Who Have Sex with Men, Atlanta, 2010–2014

|  | Partnership Type (Odds Ratio, 95% CI) |  |  |
| --- | --- | --- | --- |
|  | Main, Casual, & One-Time (n = 1397) | Main (n = 253) | Casual (n = 494) |
| **Unknown = No<sup>A</sup>** |  |  |  |
| **Absolute Age Difference** |  |  |  |
| Per 5 Years | 0.91 (0.81, 1.02) | 0.91 (0.72, 1.15) | 0.90 (0.79, 1.04) |
| **Racial Composition** |  |  |  |
| Black–Black | 1.00 | 1.00 | 1.00 |
| Black–White | 1.10 (0.50, 2.41) | 2.23 (0.43, 11.50) | 0.94 (0.37, 2.38) |
| White–White | 1.41 (0.96, 2.07) | 2.42 (1.26, 4.66) | 0.99 (0.61, 1.61) |
| **Perceived Concordant HIV Status** |  |  |  |
| Discordant/Unknown | 1.00 | 1.00 | 1.00 |
| Concordant | 0.82 (0.55, 1.21) | 1.10 (0.54, 2.26) | 0.73 (0.45, 1.19) |
| **Coital Frequency in Past 6 Months** |  |  |  |
| Per 5 Acts | 1.00 (0.95, 1.06) | 1.05 (0.97, 1.13) | 0.92 (0.84, 1.00) |
| **Partnership Agreement about Outside Sex** |  |  |  |
| No Agreement | 1.00 | 1.00 | 1.00 |
| No Outside Sex | 1.85 (1.06, 3.21) | 1.88 (0.90, 3.94) | 0.91 (0.15, 5.59) |
| Outside Sex with Conditions | 0.58 (0.28, 1.20) | 0.37 (0.12, 1.18) | 0.79 (0.28, 2.23) |
| Outside Sex without Conditions | 1.15 (0.59, 2.23) | 0.98 (0.40, 2.40) | 1.03 (0.29, 3.68) |
| **Unknown = Missing<sup>B</sup>** |  |  |  |
| **Absolute Age Difference** |  |  |  |
| Per 5 Years | 0.87 (0.76, 0.99) | 0.88 (0.69, 1.12) | 0.86 (0.73, 0.97) |
| **Racial Composition** |  |  |  |
| Black–Black | 1.00 | 1.00 | 1.00 |
| Black–White | 1.11 (0.45, 2.77) | 3.82 (0.54, 26.79) | 0.89 (0.31, 2.56) |
| White–White | 1.54 (0.99, 2.40) | 2.31 (1.11, 4.81) | 1.12 (0.63, 1.98) |
| **Perceived Concordant HIV Status** |  |  |  |
| Discordant/Unknown | 1.00 | 1.00 | 1.00 |
| Concordant | 0.83 (0.52, 1.31) | 1.03 (0.46, 2.30) | 0.73 (0.42, 1.28) |
| **Coital Frequency in Past 6 Months** |  |  |  |
| Per 5 Acts | 1.06 (0.99, 1.12) | 1.14 (1.04, 1.25) | 0.95 (0.87, 1.04) |
| **Partnership Agreement about Outside Sex** |  |  |  |
| No Agreement | 1.00 | 1.00 | 1.00 |
| No Outside Sex | 2.59 (1.40, 4.80) | 2.04 (0.86, 4.79) | 1.37 (0.21, 9.10) |
| Outside Sex with Conditions | 0.75 (0.32, 1.73) | 0.39 (0.11, 1.31) | 1.01 (0.26, 3.94) |
| Outside Sex without Conditions | 1.47 (0.73, 2.97) | 0.89 (0.32, 2.46) | 1.46 (0.41, 5.22) |

<sup>A</sup> Responses for unknown baseline degree were coded as not ongoing.
<sup>B</sup> Responses for unknown baseline degree were coded as missing and observations dropped from the model.

In Table 4, we show the result of the Poisson regression models estimating the association between confirmed degree (outcome) and baseline degree (predictor). Marginal degree distributions for both degree measures are provided in Supplemental Table 1. Models are stratified by participants who had main and casual partnerships (n = 405), any main partnerships (n = 220), or any casual partnerships (n = 299). We also ran models for the different classifications of unknown baseline status partnerships, and with adjustment for participant age and race. Baseline degree was strongly associated with confirmed degree across partnership types, for both unknown status classification methods and after demographic adjustment. Positive values of these regression coefficients indicate an increase in predicted confirmed degree with each one-unit increase in baseline degree: confirmed degree increased by a relative factor of 1.6 (*e*^*0.48*^) with each increase in baseline degree. The coefficient sizes were greater and confidence intervals wider for estimation with unknown baseline degree coded as missing as a result of greater effect size but smaller sample size.

**Table 4.** Individual-Level Poisson Regression for Confirmed Degree, by Partnership Type History, as a Function of Baseline Degree in a Study of Black and White Men Who Have Sex with Men, Atlanta, 2010–2014.

|  | Confirmed Degree<br>Coefficient (95% CI) |  |  |
| --- | --- | --- | --- |
|  | *Main or Casua*<sup>A</sup><br>(*n* = 405) | *Any Main*<sup>B</sup><br>(*n* = 220) | *Any Casual*<sup>C</sup><br>(*n* = 299) |
| **Unknown = No<sup>D</sup>** |  |  |  |
| **Unadjusted Model** |  |  |  |
| Intercept | -1.04 (-1.24, -0.85) | -1.28 (-1.65, -0.94) | -1.35 (-1.60, -1.11) |
| Baseline Degree | 0.48 (0.34, 0.56) | 0.75 (0.38, 1.13) | 0.52 (0.38, 0.65) |
| **Adjusted Model** |  |  |  |
| Intercept | -1.33 (-1.90, -0.75) | -1.73 (-2.63, -0.84) | -1.74 (-2.52, 0.97) |
| Baseline Degree | 0.46 (0.34, 0.57) | 0.79 (0.41, 1.19) | 0.53 (0.39, 0.67) |
| Age (Per 5 years) | 0.00 (-0.10, 0.10) | 0.03 (-0.13, 0.18) | 0.03 (-0.11, 0.16) |
| Race |  |  |  |
| *Black* | 0.00 | 0.00 | 0.00 |
| *White* | 0.41 (0.13, 0.70) | 0.42 (0, 0.86) | 0.34 (-0.04, 0.73) |
| **Unknown = Missing<sup>E</sup>** |  |  |  |
| **Unadjusted Model** |  |  |  |
| Intercept | -1.20 (-1.49, -0.93) | -1.55 (-2.05, -1.08) | -1.49 (-1.87, -1.13) |
| Baseline Degree | 0.48 (0.34, 0.61) | 0.97 (0.51, 1.44) | 0.56 (0.38, 0.72) |
| **Adjusted Model** |  |  |  |
| Intercept | -1.45 (-2.19, -0.71) | -1.97 (-3.01, -0.94) | -1.74 (-2.77, -0.71) |
| Baseline Degree | 0.48 (0.34, 0.63) | 1.05 (0.57, 1.55) | 0.57 (0.39, 0.74) |
| Age (Per 5 years) | -0.01 (-0.14, 0.12) | 0 (-0.18, 0.17) | 0.01 (-0.18, 0.18) |
| Race |  |  |  |
| *Black* | 0.00 | 0.00 | 0.00 |
| *White* | 0.46 (0.08, 0.85) | 0.56 (0.08, 1.06) | 0.31 (-0.19, 0.84) |

<sup>A</sup> Study participants who had any main or any casual partnerships in the 6 months prior to the baseline survey; degree is the sum of main and casual partnerships that were ongoing.
<sup>B</sup> Study participants who had any main partnerships in the 6 months prior to the baseline survey; degree is the sum of main partnerships that were ongoing.
<sup>C</sup> Study participants who had any casual partnerships in the 6 months prior to the baseline survey; degree is the sum of casual partnerships that were ongoing.
<sup>E</sup> Responses for unknown baseline degree were coded as not ongoing.
<sup>F</sup> Responses for unknown baseline degree were coded as missing and observations dropped from the model.

The bias in baseline degree for was -0.28 overall when unknown ongoing partnerships were coded as not ongoing and -0.49 when coded as missing. This bias also emerged in estimates of concurrency, or the proportion of men exhibiting a degree of 2 or more across main and casual partnerships. The proportion of participants concurrent using baseline degree only was 17.2% when unknown ongoing partnerships were coded as not ongoing and 22.0% when coded as missing. The proportion of participants concurrent based on confirmed degree was 9.6%.

The Figure shows the predicted difference between confirmed and baseline degree as a function of baseline degree from the model in which unknown baseline partnerships were classified as not ongoing. If baseline degree perfectly predicted confirmed degree, the estimates (points) would fall on the horizontal dashed line representing no difference between baseline and confirmed degrees. The plots were stratified by partnership type, with x-axis ranges limited to the range of empiric data for each type. Confirmed degree was 0.28, 0.59, and 1.25 for those with baseline degrees of 0, 1, and 2 main partnerships, respectively. The corresponding bias sizes were therefore 0.28, -0.41, and -0.75. Confirmed degree was 0.26, 0.44, 0.74, 1.24, 2.09, and 3.51 for those with baseline degree of 0 to 5 casual partnerships, respectively. The corresponding bias sizes were 0.26, -0.56, -1.26, -1.76, -1.91, and -1.49. Therefore, baseline degree overestimated confirmed degree for MSM engaged in any ongoing partnerships (degree > 0), but underestimated degree for MSM in no ongoing partnerships at baseline.

In eFigure 2, we present the model-estimated confirmed degree as a function of baseline degree rather than the bias itself, stratified by race and age group instead of partnership type. A perfect prediction of degree from baseline degree would result in the point estimates overlapping with the diagonal dashed identity line. The associations here were similar to the main text Figure with respect to baseline degree as the primary predictor of interest. However, the level of bias was greater for younger MSM (age 18–29) compared to older MSM and for black MSM compared to white MSM. For example, for MSM with a baseline degree of 2, the predicted confirmed degree was 0.65 for Black MSM aged 18–29, 0.94 for White MSM aged 18–29, 0.79 for Black MSM aged 30–39, and 1.15 for White MSM aged 30–39.

## DISCUSSION

In this study, we evaluated the potential for bias in estimating sexual network degree using only a cross-sectional (“ day-of-study”) measure by reconfirming the ongoing status of partnerships with prospective cohort data. We found high accuracy of the baseline ongoing status using this longitudinal data as the standard, with generally greater accuracy in main compared to casual partnerships but with the results dependent upon categorization of unknown status partnerships. Baseline degree overestimated true network degree when confirmed longitudinally, with the level of bias ranging from -1.91 to -0.41 for men with any ongoing partnerships. Baseline degree underestimated true degree for men with no predicted ongoing partnerships at baseline (0.26–0.28 average bias). The bias across baseline degree for main and casual partnerships was -0.28. These results held across partnership type and other covariates, but the bias was higher for younger and for black MSM.

These findings have implications for how we measure and interpret network degree and related network measures. Our study provides strong support for the need to validate network measures such as degree. Errors in the classification of the ongoing status of partnerships also impact estimation of partnership duration, since partnerships assumed to be ongoing have a censored end date, leading to a overestimation of duration if partnerships are truly not ongoing.^14,24^ From an HIV transmission perspective, overestimation of degree and duration may lead to conclusions that MSM networks are riskier than they truly are. After confirmation with longitudinal data, the fraction exhibiting concurrency was roughly half of that estimated based on baseline data alone (9.6% versus 17.2%). Although the biases in degree found here appear small, minor differences in network measures at the individual-level can cascade into large network-level outcomes like the temporal network path reachable by infectious disease pathogens.^25^

Biases in network measurements have consequences for using networks to explain HIV disparities by race in the U.S. HIV disparities research has been challenged over the past two decades to identify explanatory factors for the large racial disparities in HIV prevalence; individual-level risk factors (e.g., coital frequency and condom use) have consistently been equal or lower in the group with higher prevalence (i.e., black MSM).^6,17^ Empirical studies across multiple populations of MSM have suggested some racial differences in network degree, although the results are not consistent.^22,26,27^ We found that the level of bias in degree measurements was higher for black MSM compared to white MSM, and conversely the accuracy of baseline ongoing status was highest in white–white partnerships. We had hypothesized that the groups with higher HIV incidence (black MSM) would overestimate degree less than groups with lower incidence (white MSM), which would strengthen the role of network factors in understanding disparities.^17^ We found just the opposite: a greater overestimation of degree for black MSM. This suggests that the explanatory power of network factors in explaining disparities could be diluted.

The estimated biases support reevaluation of network measures used in epidemiologic research. Network-based mathematical modeling research in particular depends on unbiased measures of degree and duration to simulate HIV transmission dynamics.^21^ Our findings that bias was higher in casual compared to main partnerships and for MSM with larger baseline degrees suggests empiric models may have overestimated the role of these factors for explaining HIV transmission.^17,28^ Intervention models projecting the impact of new prevention strategies (such as HIV PrEP) based on network structure may also overestimate effectiveness if model parameters for degree are inaccurate.^10,19^

Network measures such as degree may be used for a number of public health activities, including STI partner notification. The goal of partner notification is to diagnose and treat (or link to treatment) partners of index patients recently diagnosed with an STI.^20^ This prevents both reinfection to the index patient by infected partners and further dissemination of infection to other partners in the sexual network.^29^ Due to funding constraints, health departments tasked with partner notification must select which partners are the highest priority to contact;^30^ to prevent repeat index infections (e.g., of curable bacterial STIs) the priority would be ongoing partnerships. Our findings suggest considerable uncertainty in defining those partnerships, which may create inefficiencies in targeting that should be considered further.

### Limitations

First, we excluded 16% of individuals and 21% of partnerships enrolled in the cohort, largely due to missing or unknown data for baseline or confirmed (M6) degree. This resulted in fewer one-time partnerships and more main/casual partnerships, which could have resulted in a higher level of estimated of bias if they had been included. Second, other factors within the longitudinal study design may have resulted in lower degree at follow-up in a way that could contribute to the observed biases: 1) participant recall issues about partnerships at baseline; and 2) participant knowledge at follow-up that the study survey is completed faster when reporting on fewer partners. Although it is impossible to know how much these alternative explanations could have affected the results, they were our primary motivation to limit the comparisons to baseline and Month 6 data and not data from subsequent follow-up visits. Finally, the study truncated data collection at the 5 most recent partners, which could result in underestimation of baseline degree; however, as eTable 1 shows, the number of participants reporting a degree of 4 or 5 was small.

### Conclusions

Our findings may have at least five implications for future network research. Our conclusions may be most applicable to MSM sexual networks that were the focus of this analysis, but could impact measurement of degree in other study populations depending upon transportability assumptions.^31^ First, if longitudinal degree data are available for other target populations, one may directly quantify potential biases using our methods. Second, if longitudinal data are not available and it is reasonable to transport our estimates, applying our estimated bias factors to other cross-sectional estimates may suffice. Third, if longitudinal data are not available but unknown ongoing status is measured, reclassifying unknown partnerships to not ongoing may reduce bias, as it did here. Fourth, if retrospective data on degree at multiple time points is available, comparison of degree over time would be a useful evaluation of measure stability. Previous studies have directly compared measures of concurrency across retrospective time points;^13,32^ this would be a useful exercise for network degree and other network measures more generally. Fifth, with any of the above four approaches to degree adjustment and evaluation above, structured sensitivity analyses, with degree ranging from that observed value to the estimated biased value, will help to understand how biased degree measures could impact the primary study outcome or clinical decision.^33^ We hope this will inform future research or public health practice activities that incorporate degree and related network measures.

## Data Availability

Primary data are not available due to IRB restrictions.

